# THE ASSOCIATION OF NUTRITIONAL STATUS ON FUNCTIONAL CAPACITY AND QUALITY OF LIFE IN CARDIAC AMYLOIDOSIS PATIENTS: AN EXPLORATORY PILOT STUDY

**DOI:** 10.64898/2026.02.27.26347247

**Authors:** Paula Aver Bretanha Ribeiro, Shana Souza Grigoletti, Priccila Zuchinali, Anne-Sophie Zenses, Valerie Fontaine, Stephania Argentin, François Tournoux

## Abstract

**Aims:** This study aimed to examine the prevalence of malnutrition and its associations with functional capacity and quality of life (QoL) in AL and ATTR cardiac amyloidosis patients.

**Methods and Results:** This cross-sectional pilot study included 29 patients with confirmed CA (14 AL, 15 ATTR). Data were collected between January 2020 and September 2021. Nutritional status was assessed using body mass index (BMI), anthropometric measures, and the Subjective Global Assessment (SGA). Functional capacity was evaluated via handgrip strength and the 6-minute walk test, while QoL was assessed using the SF-36 and Kansas City Cardiomyopathy Questionnaire. Malnutrition, as determined by SGA, was present in 62% of patients, with no significant difference between AL and ATTR subtypes. In contrast, BMI according to WHO criteria failed to identify any cases of malnutrition, highlighting its limited utility in this population. These results suggest that conventional indicators may underestimate nutritional impairment in CA. Although overall QoL and functional capacity did not differ significantly between nutritional groups, malnourished AL patients showed notably lower QoL scores compared with well-nourished peers.

**Conclusion:** Malnutrition is highly prevalent in cardiac amyloidosis and seems to particularly affect the AL subtype. These findings underscore the importance of routine nutritional screening and targeted interventions, as early identification and management of malnutrition may improve patients’ quality of life and long-term outcomes.

## 1. Introduction

Amyloidosis is a clinical disorder that is characterized by abnormal folding of proteins, leading to the formation of insoluble fibrils that accumulate within different organs and alter their structure and function. Different proteins can lead to amyloidosis, but two main forms can affect the heart ^1^: amyloid light chain (AL) amyloidosis and amyloid transthyretin-related (ATTR) amyloidosis. The prognosis of patients affected by AL or ATTR amyloidosis is predominantly correlated with the severity of cardiac involvement ^2^. Because ATTR and AL amyloidosis are systemic diseases, patients can present heart failure symptoms as well as extracardiac signs, depending on which other organs are affected. This may account for the observation that patients with cardiac amyloidosis (CA) often report, to a greater extent than most heart failure (HF) patients, a markedly reduced quality of life (QOL) and functional impairments in both physical and mental health domains ^3-5^. Among them, malnutrition and weight loss are prominent clinical features ^6^.

Malnutrition is a complex and multifactorial process that can result from reduced dietary intake, diminished absorption of macro and/or micronutrients, or increased energy expenditure^7^. Several amyloidosis patients present clinical manifestations that include nausea and/or vomiting, dysphagia, dysgeusia, diarrhea, malabsorption, anorexia, and weight loss^8-12^, which may lead to nutritional impairment. This condition can also be exacerbated by disease progression, the number of organs affected, treatment side effects, and aging. Despite this, few studies have focused on assessing nutritional status as the main outcome in patients with amyloidosis, whether of the ATTR or AL type. There is a high prevalence of malnutrition among patients with AL amyloidosis (ranging from 25% to 65%^13^), while only a few studies have assessed nutritional status in patients with ATTR cardiac amyloidosis^14-16^ (even if there might be an association between impaired nutritional status and survival^17^ in this population). Regular assessment of the nutritional status in this population could be essential in order to provide an early, effective, and appropriate tailored intervention that might improve clinical outcomes^18^. This study aimed to explore the nutritional health, functional abilities, and quality of life in patients with CAs, and to examine how these factors relate to each other.

## 2. Methods

We conducted a cross-sectional observational study at the University of Montreal Health Centre (Centre Hospitalier de l’Université de Montréal, CHUM) in Montreal, Canada. We collected data from January 2020 to March 2025 and invited all patients in our Cardiac Amyloidosis database to participate. The study protocol was approved by the Research Ethics Committee of CHUM (2020-8705), and written informed consent from each patient was obtained.

### 2.1 Participants

Patients were eligible if they were diagnosed with CA according to current Canadian guidelines and were at least 18 years old. Participants who presented physical or mental conditions that prevented them from consenting or performing the evaluations were excluded. Demographic, imaging, laboratory, and clinical data were collected from the patient’s electronic medical records.

### 2.2 Nutritional Status

A dietitian was responsible for assessing the nutritional status of each enrolled patient. To determine the nutritional status, anthropometry (weight, height, mid-upper arm circumference, and triceps skinfold thickness), the body mass index (BMI), and the Subjective Global Assessment (SGA) were used^19^.

SGA is a validated tool that uses objective and subjective measures to classify the severity of malnutrition. It is a multi-component instrument that considers weight, weight change, dietary intake and change, gastrointestinal symptoms (duration and frequency), and functionality, as well as physical examination. Patients were categorized as SGA-A if “no signs of malnutrition”; SGA-B if “mild or moderate malnutrition”; and SGA-C if “severe malnutrition”. For the purposes of this analysis, participants were classified into two categories: well-nourished (SGA-A) and malnourished (SGA-B and C).

Anthropometric measures, including weight, height, mid-upper arm circumference (MAC), and triceps skinfold thickness (TSF), were assessed according to standard procedures ^20^, and used to calculate mid-arm muscle circumference (MAMC) and BMI. Patients were classified according to BMI using the World Health Organization classification or, if aged 65 years and older as: underweight (<22 kg/m^2)^, normal weight (22-27 kg/m^2^), or overweight/obese (>27 kg/m^2^)^20,21^. Muscle depletion was defined as a MAMC below the 10th percentile of a reference population adjusted for age and gender^22^.

### 2.3 Functional capacity and physical activity assessments

To assess performance status, the Eastern Cooperative Oncology Group (ECOG) performance status scale was used^23^. Handgrip strength was measured using a hydraulic hand dynamometer (Jamar®, Sammons Preston, Bolingbrook, IL, USA). For this test, patients were in a seated position, with their elbow flexed at 90°, and their forearm and wrist in a neutral position. Participants were instructed to perform three maximal isometric contractions. Patients took recovery pauses between measurements. The mean of three attempts with both hands was recorded. A cut-off point of 32.2 Kgf was used to define low muscle strength^24^.

Submaximal functional capacity was assessed with the 6-minute walk test (6MWT), which was performed according to international guidelines^25,26^. Briefly, patients were instructed to cover the greatest distance possible at their own pace in a 20 m corridor during the 6 minutes. Patients were allowed to use their walking aids during the tests.

Finally, the level of physical activity was determined using the International Physical Activity Questionnaire (IPAQ) for walking and leisure time questions. Patients were considered active if time in physical activity was >90min/week^27^.

### 2.4 Quality of life assessments

General health-related Quality of life was assessed by the 36-item Short-Form General Health Survey (SF-36) with a 4-week recall. The SF-36 assesses eight specific domains of functional health and well-being: Physical Functioning (PF), Role-Physical (RP; role limitations due to physical problems), Bodily Pain (BP), General Health Perceptions (GH), Vitality (VT), Social Functioning (SF), Role -Emotional (RE; role limitations due to emotional problems), and Mental Health (MH). The physical component summary (PCS) is derived from PF, RP, BP, and GH, and the mental component summary (MCS) is derived from VT, SF, RE, and MH. All scale and summary scores were calculated using the developer’s scoring algorithm^28^.

The Kansas City Cardiomyopathy Questionnaire (KCCQ), with twelve items, was used to assess disease-specific quality of life, within a 2-week recall period. Questions targeting four domains: physical limitation, symptom frequency, social limitation, and quality of life. Scores range from 0 to 100, with lower scores reflecting a poorer quality of life, and were calculated using the developer’s scoring algorithm^29^.

### 2.5 Statistical Analyses

Results are expressed as means and SD, or as the number of cases and proportions (%), total and according to nutritional status (well-nourished and malnourished). Fisher’s exact test was used to compare categorical and nominal results, and T-test for independent samples was used to compare groups. All data were collected and stored at the institutional REDCAP, a secure, web-based software platform designed to support data capture for research studies^30,31^. Statistical significance was set at an alpha level of.05, and all analyses were performed using SPSS version 29 (Chicago, IL, USA).

## 3. Results

Twenty-nine patients were enrolled in the study. Patients’ clinical characteristics in the two groups were comparable for most general characteristics, such as age, comorbidities, and blood pressure, data presented in Table 1. The mean age was 74. ±11 years, and 79% were male. The study population included 52% of patients with confirmed ATTR-CA and 48% with AL-CA. The kidney was the second most frequently affected organ in our patients (14%). Most patients (52%) were in NYHA class II, and 11% had reduced left ventricular ejection fraction.

**Table 1:**
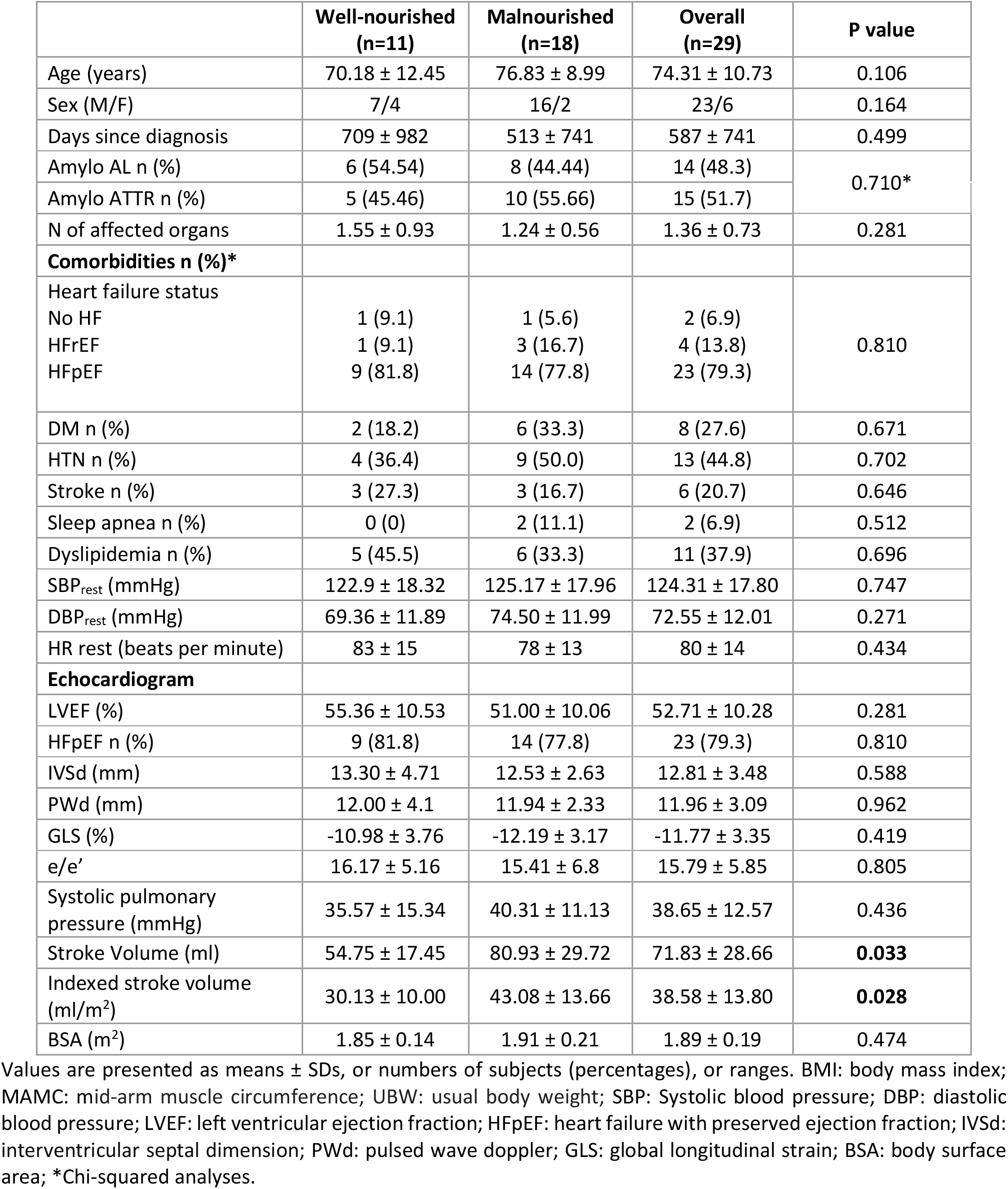
Patients’ clinical characteristics according to nutritional status.

### Nutritional Status

The SGA identified 62% (n=18) of the patients with malnutrition. Among them, 17 were categorized as SGA-B (mild or moderate malnutrition) and one as SGA-C (severe malnutrition). Fifty-five percent of the patients described in the malnourished group presented AL amyloidosis. A total of sixteen patients reported unintentional WL in the previous 6 months at some level (details are in Table 2). The dexamethasone treatment (total n=12) was tested between SGA groups, and it did show differences (p=1.000).

**Table 2:** Nutritional characteristics according to SGA nutritional status.

|  | **Well-nourished**<br>**(n=11)** | **Malnourished**<br>**(n=18)** | **Overall**<br>**(n=29)** | **P value** |
| --- | --- | --- | --- | --- |
| BMI (kg/m<sup>2</sup>) | 29.44 ± 6.68 | 25.31 ± 3.57 | 26.88 ± 5.27 | **0.038** |
| Underweight BMI (WHO) | 0 (0) | 0 (0) | 0 (0) | NA |
| Underweight BMI age-adapted | 0 (0) | 4 (22.2) | 4 (13.8) | 0.268 |
| Overweight + Obesity n (%) age-adapted | 7 (63.6) | 3 (16.7) | 10 (34.5) | **0.017** |
| Overweight + Obesity n (%) **(WHO)** | 8 (72.7) | 10 (55.6) | 18 (62.1) | 0.449 |
| **SGA items** |  |  |  |  |
| **Inadequate dietary intake (n=28)** | 0 (0) | 9 (50) | 9 (32.1) | **0.010** |
| **GI symptoms** |  |  |  |  |
| Nausea n (%) | 2 (18.2) | 4 (22.2) | 6 (20.7) | 1.000 |
| Vomiting n (%) | 0 (0) | 2 (11.1) | 2 (6.9) | 0.512 |
| Diarrhea n (%) | 0 (0) | 6 (33.3) | 6 (20.7) | **0.058** |
| Dysphagia n (%) | 1 (9.1) | 7 (38.9) | 8 (27.6) | 0.110 |
| Anorexia n (%) | 0 (0) | 1 (5.6) | 1 (3.4) | 1.000 |
| Early satiety n (%) | 1 (9.1) | 9 (50) | 19 (34.5) | **0.044** |
| Constipation n (%) | 3 (27.3) | 3 (16.7) | 6 (20.7) | 0.646 |
| **Weight change** |  |  |  |  |
| Self-reported weight loss (kg) | 0.29 ± 0.65 | 4.66 ± 3.10 | 3.04 ± 3.27 | **<0.001** |
| Self-reported weight loss / n (%) | 2 (18.2) | 14 (77.8) | 16 (55.2) |  |
| Weight loss <5% UBW | 2 (18.2) | 3 (16.7) | 5 (17.2) | **0.007** |
| Weight loss 5 – 10 % UBW | 0 (0) | 7 (38.9) | 7 (24.1) |  |
| Weight loss >10% UBW | 0 (0) | 4 (22.2) | 4 (13.8) |  |

Values are presented as means ± SDs, or numbers of subjects (percentages), or ranges. BMI: body mass index; UBW: usual body weight; \* 2 missing values.

According to the BMI age-adapted, only 14% (n=4) of the subjects were classified as underweight, 52% (n=15) as normal weight, and 34% (n=10) as overweight or obese. However, among the malnourished patients, 17% (n=3) were identified as overweight or obese based on BMI values, highlighting that malnutrition can be subtle and not readily apparent in some cases. Of the total 18 malnourished according to SGA, only 4 were considered underweight according to BMI age-adapted (Kappa = 0.178). When analyzing the patient’s BMI according to the WHO classification, data revealed that 56% of the malnourished patients (as identified by SGA), were overweight or obese. Furthermore, according to the same BMI classification, none of the patients were classified as underweight; therefore, Kappa could not be calculated.

Nutritional depletion was detected using MAMC. Results demonstrated values below the sex- and age-matched 10th percentile in 10 patients (34.5%), regardless of group (see Table 1 for details). Overall, there were no significant differences between groups (Well-nourished vs. Malnourished groups) for MAMC.

### Functional Capacity, Quality of Life, and Physical Activity

Data on functional capacity and QOL stratified by SGA categories and amyloidosis type are reported in Table 3. In terms of functional capacity, 18 patients (62%) presented handgrip weakness. More than ¾ of our malnourishment patients had handgrip weakness (14 out of 18; coexistent comorbidities). ECOG demonstrated that a total of 14 patients had a score ≥2 (i.e., unable to carry out any work activities), among which 11 were malnourished. Overall, the ECOG performance status scale was 0, 1, 2, and 3 in 2, 13, 10, and 4 patients, respectively.

**Table 3:**
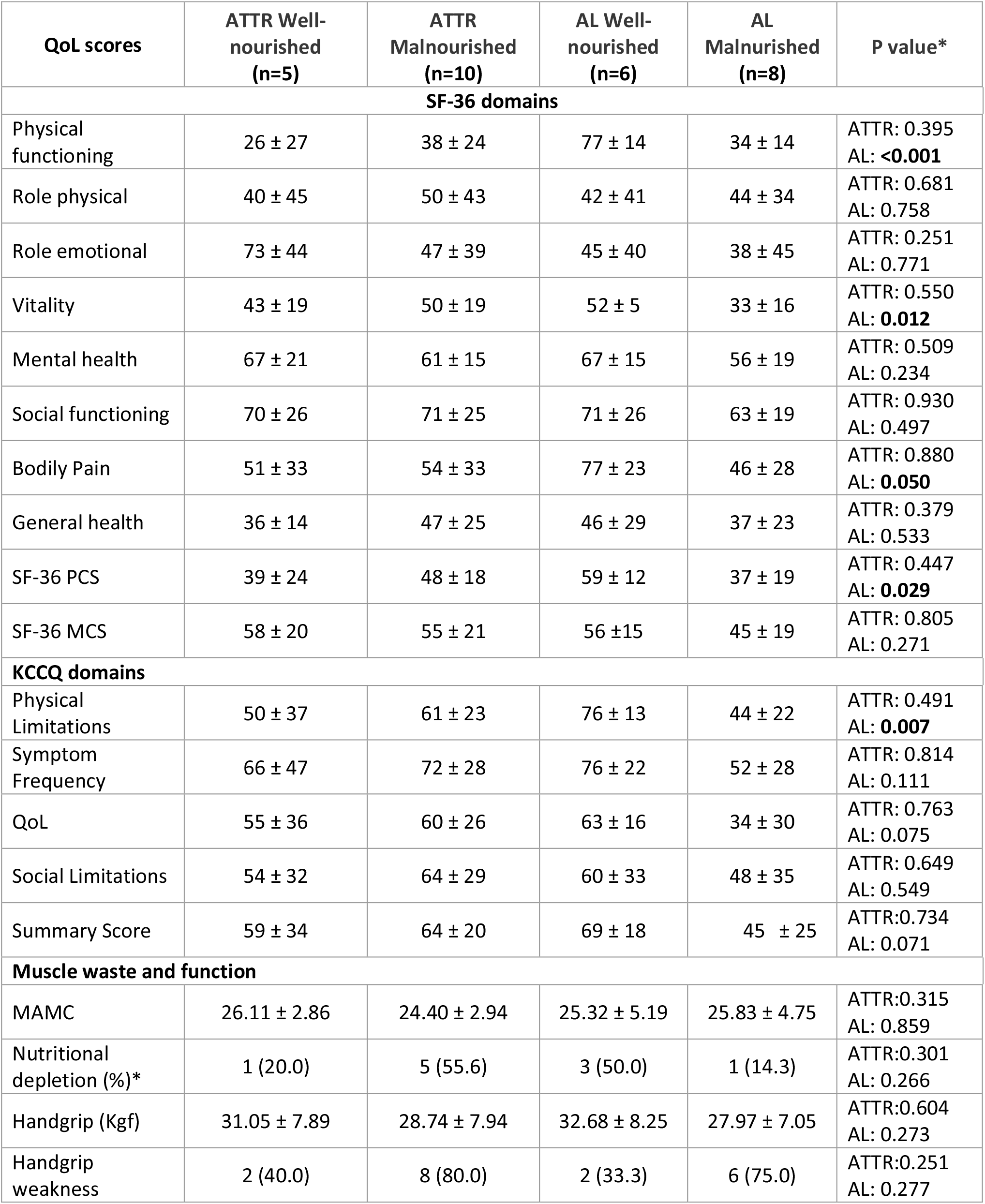
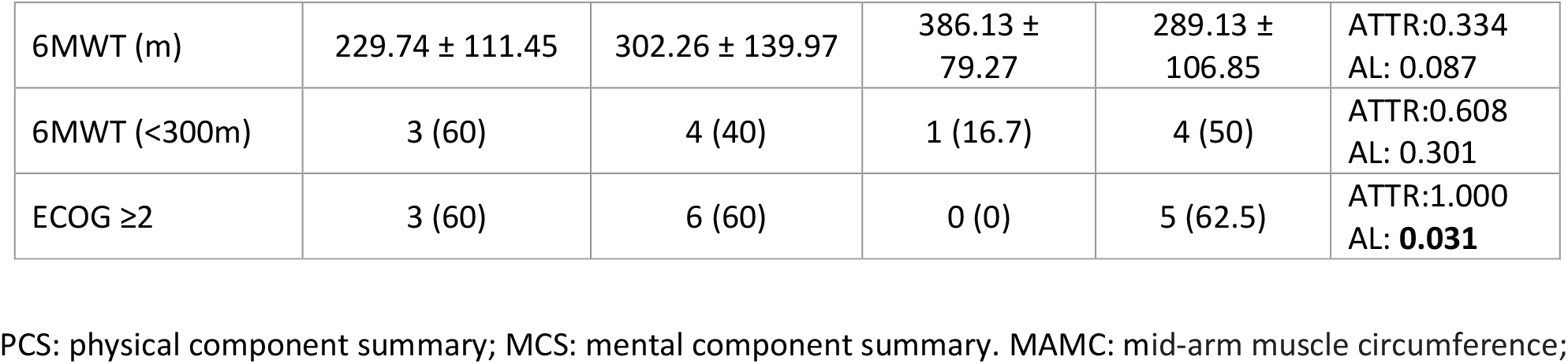
Quality of life and functional capacity according to nutritional status and amyloidosis subtype.

| QoL scores | ATTR Well-nourished (n=5) | ATTR Malnourished (n=10) | AL Well-nourished (n=6) | AL Malnourished (n=8) | P value* |
| --- | --- | --- | --- | --- | --- |
| **SF-36 domains** |  |  |  |  |  |
| Physical functioning | 26 ± 27 | 38 ± 24 | 77 ± 14 | 34 ± 14 | ATTR: 0.395<br>AL: **<0.001** |
| Role physical | 40 ± 45 | 50 ± 43 | 42 ± 41 | 44 ± 34 | ATTR: 0.681<br>AL: 0.758 |
| Role emotional | 73 ± 44 | 47 ± 39 | 45 ± 40 | 38 ± 45 | ATTR: 0.251<br>AL: 0.771 |
| Vitality | 43 ± 19 | 50 ± 19 | 52 ± 5 | 33 ± 16 | ATTR: 0.550<br>AL: **0.012** |
| Mental health | 67 ± 21 | 61 ± 15 | 67 ± 15 | 56 ± 19 | ATTR: 0.509<br>AL: 0.234 |
| Social functioning | 70 ± 26 | 71 ± 25 | 71 ± 26 | 63 ± 19 | ATTR: 0.930<br>AL: 0.497 |
| Bodily Pain | 51 ± 33 | 54 ± 33 | 77 ± 23 | 46 ± 28 | ATTR: 0.880<br>AL: **0.050** |
| General health | 36 ± 14 | 47 ± 25 | 46 ± 29 | 37 ± 23 | ATTR: 0.379<br>AL: 0.533 |
| SF-36 PCS | 39 ± 24 | 48 ± 18 | 59 ± 12 | 37 ± 19 | ATTR: 0.447<br>AL: **0.029** |
| SF-36 MCS | 58 ± 20 | 55 ± 21 | 56 ± 15 | 45 ± 19 | ATTR: 0.805<br>AL: 0.271 |
| **KCCQ domains** |  |  |  |  |  |
| Physical Limitations | 50 ± 37 | 61 ± 23 | 76 ± 13 | 44 ± 22 | ATTR: 0.491<br>AL: **0.007** |
| Symptom Frequency | 66 ± 47 | 72 ± 28 | 76 ± 22 | 52 ± 28 | ATTR: 0.814<br>AL: 0.111 |
| QoL | 55 ± 36 | 60 ± 26 | 63 ± 16 | 34 ± 30 | ATTR: 0.763<br>AL: 0.075 |
| Social Limitations | 54 ± 32 | 64 ± 29 | 60 ± 33 | 48 ± 35 | ATTR: 0.649<br>AL: 0.549 |
| Summary Score | 59 ± 34 | 64 ± 20 | 69 ± 18 | 45 ± 25 | ATTR: 0.734<br>AL: 0.071 |
| **Muscle waste and function** |  |  |  |  |  |
| MAMC | 26.11 ± 2.86 | 24.40 ± 2.94 | 25.32 ± 5.19 | 25.83 ± 4.75 | ATTR: 0.315<br>AL: 0.859 |
| Nutritional depletion (%)* | 1 (20.0) | 5 (55.6) | 3 (50.0) | 1 (14.3) | ATTR: 0.301<br>AL: 0.266 |
| Handgrip (Kgf) | 31.05 ± 7.89 | 28.74 ± 7.94 | 32.68 ± 8.25 | 27.97 ± 7.05 | ATTR: 0.604<br>AL: 0.273 |
| Handgrip weakness | 2 (40.0) | 8 (80.0) | 2 (33.3) | 6 (75.0) | ATTR: 0.251<br>AL: 0.277 |
| 6MWT (m) | 229.74 ± 111.45 | 302.26 ± 139.97 | 386.13 ± 79.27 | 289.13 ± 106.85 | ATTR:0.334<br>AL: 0.087 |
| 6MWT (<300m) | 3 (60) | 4 (40) | 1 (16.7) | 4 (50) | ATTR:0.608<br>AL: 0.301 |
| ECOG ≥2 | 3 (60) | 6 (60) | 0 (0) | 5 (62.5) | ATTR:1.000<br>AL: **0.031** |

PCS: physical component summary; MCS: mental component summary. MAMC: mid-arm muscle circumference;

For QoL scores, well-nourished AL patients had consistently higher average scores, and five of them were statistically significant – details in Table 3. Overall, physical activity did not differ between well-nourished and malnourished patients, and the same proportion of patients were considered active in both groups (p=1.000).

## 4. Discussion

Findings from our pilot study indicate that poor nutritional status, assessed by the Subjective Global Assessment (SGA), is common in patients with cardiac amyloidosis, affecting 62% of our cohort. Notably, none of the malnourished patients were classified as underweight according to standard BMI criteria, highlighting the limitations of BMI as a screening tool. Although we were able to establish a significant association between impaired nutritional status and quality of life only for AL patients —likely due to limited statistical power—our findings suggest that incorporating nutritional and functional assessments may enhance disease management in this population. Overall, these results underscore the importance of systematic malnutrition screening, which warrants further investigation in larger studies.

### Nutritional Status

In concordance with our results, Sattianayagam et al., who used Patient–Generated Subjective Global Assessment (PG-SGA) score to assess the nutritional status of the patients, showed that 65% of patients with amyloidosis had malnutrition (score ≥4)^32^. Caccialanza et al. have also found similar results by assessing malnutrition according to the phase angle derived from bioelectrical impedance, an objective measurement technique. Their study reported malnutrition in 52% of patients with AL amyloidosis.

When considering only WL and/or BMI, the proportion of amyloidosis patients identified as malnourished decreases significantly. Our results showed that only 7% reported WL >10% of their usual body weight, and according to BMI, only 14% of the patients were classified as underweight. Caccialanza found that 22% of the patients had moderate malnutrition (presence of one parameter: BMI< 22 kg/m2 or WL≥10%) and 8% had severe malnutrition (both parameters combined: BMI< 22 kg/m2 and WL≥10%)^33^. In another study of the same Italian group, it was reported that 25% of patients had a BMI< 22 kg/m2, which the author indicated as the cut-off criterion for malnutrition ^34^.

Weight loss or BMI have been widely used in studies as indicators of nutritional status^13,14^, due to their accessibility and easy implementation in clinical practice. However, these are not the ideal endpoints due to high variability and misclassification (i.e., the high prevalence of malnutrition in overweight and obese patients, fluid balance, and muscle mass content), especially in cardiac amyloidosis patients. As demonstrated in our study, BMI should be used as a component of a comprehensive malnutrition screening, rather than relying on it as the sole criterion for malnutrition.

### Functional Capacity and Quality of Life

To our knowledge, no other study explored the association between nutritional status with functional capacity in patients with amyloidosis. Although preliminary, our results demonstrated that 62% of our patients had malnourishment, and among those, 78% co-existing muscle weakness. Also, the 6MWT distances were low^35^, and overall equivalent to NYHA class II^36^. Despite limited statistical power, several SF-36 physical domain scores were significantly lower in malnourished AL patients.

Previous studies also demonstrated that malnutrition could be associated with reduced quality of life. Sattianayagam et al.^32^ showed that impaired nutritional status evaluated by PG-SGA was associated with lower QoL scores using the EORTC QLQ-C30 questionnaire in AL amyloidosis (p< 0.001; n=110). Another study reported that for each kg of WL in the last six months, the summary of the mental component (SF-36 questionnaire) was reduced by 0.47 (95%CI 0.18-0.75, p= 0.002). As well, the above-mentioned study observed that the lower phase angle value (PhA) was associated with lower Physical Health Composite Scale (PCS) scores (p=0.015)^37^.

Taken together, our findings suggest that we are dealing with a complex and clinically frail population, especially the AL patients. Although malnutrition is a well-recognized predictor of poor outcomes in many chronic and progressive diseases^15,38^, it remains frequently underdiagnosed and underestimated in the context of cardiac amyloidosis. This underscores the urgent need for standardized and accurate tools to assess nutritional status in this population.

## Limitations

Our study has several limitations. It was a single-center, observational study with a relatively small sample size, which may have limited the statistical power of our analyses and warrants cautious interpretation of the results. Additionally, the inclusion of patients at varying stages of disease progression likely contributed to population heterogeneity, potentially affecting between-group comparisons. While we included all eligible patients from our clinic, the rarity of cardiac amyloidosis poses an inherent challenge to recruitment. Lastly, in the absence of disease-specific reference values for physical function assessment, we relied on thresholds derived from other populations—such as heart failure (e.g., handgrip weakness, and 6MWT) and oncology (e.g., ECOG performance status)—which may limit the specificity of our findings.

## Conclusion

Overall, impaired nutritional status has a high prevalence in our population and seems to impact quality of life, especially our AL patients. BMI failed to identify patients with malnutrition. These results reinforce the importance of systemic and standardized nutritional assessments in patients with amyloidosis. Accurate and early identification of malnutrition, through a thorough evaluation of nutritional status, will allow adequate nutritional support and may enhance the prognosis of these patients. Hence, more collaborative multi-centered studies need to be conducted to further clarify the impact of nutritional status and nutritional interventions on long-term outcomes in this population.

## Data Availability

All data produced in the present study are available upon reasonable request to the authors

## Acknowledgments

The authors would like to thank all patients who participated in this study. We would also like to thank the staff of the Cardiology Unit.

## Author Contributions

Conceptualization and design SSG, PABR, FT; Data collection SSG; Data analysis PABR, PZ; Funding acquisition SSG, PABR, FT; Writing original draft SSG; Writing – review and editing PZ, PABR, FT; Study supervision PABR and FT.

## Funding support

Scholarship from Fonds de recherche du Quebec - Nature et Technologies (FRQNT) for SSG. FT is supported by a salary grant from Fonds de recherche du Québec – Santé. This research did not receive any other specific grant from funding agencies in the public, commercial, or not-for-profit sectors.

## Conflict of interest

None of the authors has any financial or personal relationships with other people or organizations that could inappropriately influence this work.

## Notes

### Competing Interest Statement

The authors have declared no competing interest.

### Funding Statement

This study did not receive any funding

### Author Declarations

Comite d ethique a la recherche du Centre de Recherche du CHUM

## References

1. Gertz MA, Dispenzieri A, Sher T. Pathophysiology and treatment of cardiac amyloidosis. Nat Rev Cardiol. Feb 2015;12(2):91–102. doi:10.1038/nrcardio.2014.165

2. Falk RH, Alexander KM, Liao R, Dorbala S. AL (Light-Chain) Cardiac Amyloidosis: A Review of Diagnosis and Therapy. J Am Coll Cardiol. Sep 20 2016;68(12):1323–41. doi:10.1016/j.jacc.2016.06.053

3. Sanchorawala V, McCausland KL, White MK, et al. A longitudinal evaluation of health-related quality of life in patients with AL amyloidosis: associations with health outcomes over time. Br J Haematol. Nov 2017;179(3):461–470. doi:10.1111/bjh.14889

4. Wixner J, Mundayat R, Karayal ON, Anan I, Karling P, Suhr OB. THAOS: gastrointestinal manifestations of transthyretin amyloidosis-common complications of a rare disease. Orphanet journal of rare diseases 2014;9(1):1–9.

5. Lane T, Fontana M, Martinez-Naharro A, et al. Natural History, Quality of Life, and Outcome in Cardiac Transthyretin Amyloidosis. Circulation. Jul 2 2019;140(1):16–26. doi:10.1161/CIRCULATIONAHA.118.038169

6. Dubrey SW, Cha K, Anderson J, et al. The clinical features of immunoglobulin light-chain (AL) amyloidosis with heart involvement. QJM. Feb 1998;91(2):141–57. doi:10.1093/qjmed/91.2.141

7. Saunders J, Smith T. Malnutrition: causes and consequences. Clin Med (Lond). Dec 2010;10(6):624–7. doi:10.7861/clinmedicine.10-6-624

8. Kyle RA, Gertz MA. Primary systemic amyloidosis: clinical and laboratory features in 474 cases. [Sheboygan, Wis.]: Grune & Stratton,[c1964-; 1995:45–59.

9. Marinone MG, Marinone MG, Merlini G. Reduced taste perception in AL amyloidosis. A frequently unnoticed sensory impairment. Haematologica. Mar-Apr 1996;81(2):110–5.

10. Hayman SR, Lacy MQ, Kyle RA, Gertz MA. Primary systemic amyloidosis: a cause of malabsorption syndrome. Am J Med. Nov 2001;111(7):535–40. doi:10.1016/s0002-9343(01)00919-6

11. Park MA, Mueller PS, Kyle RA, Larson DR, Plevak MF, Gertz MA. Primary (AL) hepatic amyloidosis: clinical features and natural history in 98 patients. Medicine (Baltimore). Sep 2003;82(5):291–8. doi:10.1097/01.md.0000091183.93122.c7

12. Lim AY, Lee JH, Jung KS, et al. Clinical features and outcomes of systemic amyloidosis with gastrointestinal involvement: a single-center experience. Korean J Intern Med. Jul 2015;30(4):496–505. doi:10.3904/kjim.2015.30.4.496

13. Grigoletti SS, Zuchinali P, Lemieux-Blanchard E, et al. Focused review on nutritional status of patients with immunoglobulin light chain amyloidosis. Curr Probl Cancer. Jun 2022;46(3):100833. doi:10.1016/j.currproblcancer.2021.100833

14. Driggin E, Helmke S, De Los Santos J, et al. Markers of nutritional status and inflammation in transthyretin cardiac amyloidosis: association with outcomes and the clinical phenotype. Amyloid. Jun 2020;27(2):73–80. doi:10.1080/13506129.2019.1698417

15. Fumagalli C, Zampieri M, Presta R, et al. Prognostic Value of Malnutrition, Frailty, and Physical Performance in Transthyretin Cardiac Amyloidosis: Insights From a Prospective Multicenter Cohort Study. Circ Heart Fail. Jul 2 2025:e012777. doi:10.1161/circheartfailure.125.012777

16. Suhr O, Danielsson Å, Holmgren G, Steen L. Malnutrition and gastrointestinal dysfunction as prognostic factors for survival in familial amyloidotic polyneuropathy. Journal of internal medicine 1994;235(5):479–485.

17. Caccialanza R, Palladini G, Klersy C, et al. Nutritional status of outpatients with systemic immunoglobulin light-chain amyloidosis. The American journal of clinical nutrition. 2006;83(2):350–354.

18. Caccialanza R, Palladini G, Cereda E, et al. Nutritional counseling improves quality of life and preserves body weight in systemic immunoglobulin light-chain (AL) amyloidosis. Nutrition. Oct 2015;31(10):1228–34. doi:10.1016/j.nut.2015.04.011

19. Detsky A, McLaughlin J, Baker J, et al. What is Subjective Global Assessment of Nutritional Status?. JPEN J Parenter Enter Nutr. 1987 1987;11(1):8–13.

20. Frisancho AR. New standards of weight and body composition by frame size and height for assessment of nutritional status of adults and the elderly. Am J Clin Nutr. Oct 1984;40(4):808–19. doi:10.1093/ajcn/40.4.808

21. Lipschitz DA. Screening for nutritional status in the elderly. Primary Care: Clinics in Office Practice. 1994;21(1):55–67.

22. Bishop CW, Bowen P, Ritchey S. Norms for nutritional assessment of American adults by upper arm anthropometry. The American journal of clinical Nutrition. 1981;34(11):2530–2539.

23. Oken MM, Creech RH, Tormey DC, et al. Toxicity and response criteria of the Eastern Cooperative Oncology Group. Am J Clin Oncol. Dec 1982;5(6):649–55.

24. Izawa KP, Watanabe S, Osada N, et al. Handgrip strength as a predictor of prognosis in Japanese patients with congestive heart failure. European Journal of Preventive Cardiology. 2009;16(1):21–27.

25. Witham MD, Argo IS, Johnston DW, Struthers AD, McMurdo ME. Predictors of exercise capacity and everyday activity in older heart failure patients. Eur J Heart Fail. Mar 2006;8(2):203–7. doi:10.1016/j.ejheart.2005.03.008

26. Bellet RN, Adams L, Morris NR. The 6-minute walk test in outpatient cardiac rehabilitation: validity, reliability and responsiveness--a systematic review. Physiotherapy. Dec 2012;98(4):277–86. doi:10.1016/j.physio.2011.11.003

27. Craig CL, Marshall AL, Sjostrom M, et al. International physical activity questionnaire: 12-country reliability and validity. Med Sci Sports Exerc. Aug 2003;35(8):1381–95. doi:10.1249/01.MSS.0000078924.61453.FB

28. Maruish ME. User’s manual for the SF-36v2 Health Survey. Quality Metric Incorporated; 2011.

29. Green CP, Porter CB, Bresnahan DR, Spertus JA. Development and evaluation of the Kansas City Cardiomyopathy Questionnaire: a new health status measure for heart failure. J Am Coll Cardiol. Apr 2000;35(5):1245–55. doi:10.1016/s0735-1097(00)00531-3

30. Harris PA, Taylor R, Minor BL, et al. The REDCap consortium: building an international community of software platform partners. Journal of biomedical informatics. 2019;95:103208.

31. Harris PA, Taylor R, Thielke R, Payne J, Gonzalez N, Conde JG. Research electronic data capture (REDCap)—a metadata-driven methodology and workflow process for providing translational research informatics support. Journal of biomedical informatics 2009;42(2):377–381.

32. Sattianayagam PT, Lane T, Fox Z, et al. A prospective study of nutritional status in immunoglobulin light chain amyloidosis. Haematologica. Jan 2013;98(1):136–40. doi:10.3324/haematol.2012.070359

33. Caccialanza R, Palladini G, Klersy C, et al. Malnutrition at Diagnosis Predicts Mortality in Patients With Systemic Immunoglobulin Light-Chain Amyloidosis Independently of Cardiac Stage and Response to Treatment. Journal of Parenteral and Enteral Nutrition. 2014;38(7):891–894.

34. Caccialanza R, Palladini G, Klersy C, et al. Nutritional status independently affects quality of life of patients with systemic immunoglobulin light-chain (AL) amyloidosis. Ann Hematol. Mar 2012;91(3):399–406. doi:10.1007/s00277-011-1309-x

35. Casanova C, Celli BR, Barria P, et al. The 6-min walk distance in healthy subjects: reference standards from seven countries. Eur Respir J. Jan 2011;37(1):150–6. doi:10.1183/09031936.00194909

36. Yap J, Lim FY, Gao F, Teo LL, Lam CS, Yeo KK. Correlation of the New York Heart Association Classification and the 6-Minute Walk Distance: A Systematic Review. Clin Cardiol. Oct 2015;38(10):621–8. doi:10.1002/clc.22468

37. Caccialanza R, Cereda E, Klersy C, et al. Bioelectrical impedance vector analysis-derived phase angle predicts survival in patients with systemic immunoglobulin light-chain amyloidosis. Amyloid. Sep 2020;27(3):168–173. doi:10.1080/13506129.2020.1737004

38. Ruiz AJ, Buitrago G, Rodriguez N, et al. Clinical and economic outcomes associated with malnutrition in hospitalized patients. Clin Nutr. Jun 2019;38(3):1310–1316. doi:10.1016/j.clnu.2018.05.016

